# Predictive model for BNT162b2 vaccine response in cancer patients based on cytokines and growth factors

**DOI:** 10.1101/2022.09.25.22280267

**Authors:** Angelina Konnova, Fien HR De Winter, Akshita Gupta, Lise Verbruggen, An Hotterbeekx, Matilda Berkell, Laure-Anne Teuwen, Greetje Vanhoutte, Bart Peeters, Silke Raats, Isolde Van der Massen, Sven De Keersmaecker, Yana Debie, Manon Huizing, Pieter Pannus, Kristof Y Neven, Kevin K Ariën, Geert A. Martens, Marc Van Den Bulcke, Ella Roelant, Isabelle Desombere, Sébastien Anguille, Zwi Berneman, Maria E Goossens, Herman Goossens, Surbhi Malhotra-Kumar, Evelina Taconelli, Timon Vandamme, Marc Peeters, Peter van Dam, Samir Kumar-Singh

## Abstract

**Background:** Patients with cancer, especially haematological cancer, are at increased risk for breakthrough COVID-19 infection. However, so far, a predictive biomarker that can assess compromised vaccine-induced anti-SARS-CoV-2 immunity in cancer patients has not been proposed.

**Methods:** Here, we employed machine learning approaches to identify a biomarker signature based on blood cytokine and growth factors linked to vaccine response from 199 cancer patients receiving BNT162b2 vaccine.

**Results:** We show that C-reactive protein (CRP; general marker of inflammation), interleukin (IL)-15 (a pro-inflammatory cytokine), IL-18 (interferon-gamma inducing factor), and placental growth factor (an angiogenic cytokine) can correctly classify patients with a diminished vaccine response assessed at day 49 with >80% accuracy. Amongst these, CRP showed the highest predictive value for poor response to vaccine administration. Importantly, this unique signature of vaccine response was present at different studied timepoints both before and after vaccination and was not majorly affected by different anti-cancer treatments.

**Conclusion:** While we propose a blood-based signature of cytokines and growth factors that can be employed in identifying cancer patients at continued risk of COVID-19, our data also importantly suggest that such a signature could reflect the inherent make-up of some cancer patients who are also refractive to immunotherapy.

## Introduction

The field of vaccination against infectious disease has witnessed rapid advances of technology throughout the COVID-19 pandemic, including the development of various anti-SARS-CoV-2 vaccines, such as mRNA and vector vaccines. The BNT162b2 mRNA COVID-19 vaccine elicits a range of immunological responses, especially a strong anti-SARS-CoV-2 IgG response in healthy individuals, although it starts waning after approximately 6 months [1-3]. Since the mechanisms determining the quality and quantity of these responses are largely uncharacterised, this leads to concerns about the efficiency of these vaccines in vulnerable populations including patients with solid or haematological malignancies [4]. This is of special concern in cancer patients, especially under active antineoplastic treatments. For example, several studies have shown that vaccine responses are compromised in patients with haematological malignancies under B cell depleting rituximab treatment, or with solid tumours receiving different chemotherapies [5-11]. In support of these observations, we reported reduced humoral anti-SARS-CoV-2 immune responses after BNT162b2 two-dose vaccination in several but not all cancer patients under antineoplastic treatment [5]. On the other hand, only a limited number of studies are available reporting biomarkers of protection against SARS-CoV-2 infection generated by anti-SARS-CoV-2 vaccines in the general population [12, 13] and none is currently available for cancer patients where such prognostic marker can distinguish populations that are in urgent need for additional preventive options [14, 15].

Cytokine, chemokines, and certain growth factors (CCGs) are important players in the regulation of inflammation and immunity, and consequentially are strongly linked to the initiation and progression of cancers [16-19]. We recently demonstrated significant alterations in levels of several CCGs in blood of cancer patients including, but not limited to, CCGs that play an important role in the adaptive immune response in antigen presentation and/or T-helper and B cell functions [19]. Moreover, several studies have reported cytokine alterations in patients vaccinated for viruses, such as influenza [20, 21] or SARS-CoV-2 [12, 22-24]. However, again, very little data exist for CCG alterations occurring as a response to SARS-CoV-2 mRNA vaccines in cancer patients [24].

## Material and Methods

### Patient Population and study design

A prospective, longitudinal, multi-cohort trial was initiated on February 15, 2021, in the Multidisciplinary Oncological Center Antwerp (MOCA), Antwerp University Hospital, Belgium, as described[5]. Briefly, study participants aged 18 years or older with a life expectancy of at least six months were recruited. Pregnant or breastfeeding women and patients with an immunodeficiency unrelated to cancer treatment were not included. All study participants provided written informed consent. A total of 200 cancer patients recruited in this study received at least one dose of the BNT162b2 vaccine. One patient withdrew after the primer dose and was excluded from the study. CCGs before and after the primer dose were measured for 199 patients, including 158 patients with a solid tumour and 41 patients with a haematological malignancy (**Supplementary Table 1**). From these 199 patients, 187 patients received a booster dose after 21 (± 2) days after primer dose following the study protocol. Nine patients received a delayed booster dose from 24 to 37 days due to an active SARS-CoV-2-CoV-2 infection or cancer treatment-related complications[5]. The study was approved by the local ethics committee and was executed in accordance with Good Clinical Practice and the Declaration of Helsinki (ICH GCP E6(R2)). The regulatory sponsor was the Antwerp University Hospital (EudraCT number 2021-000300-38).

The majority of the population consisted of patients with breast malignancies (52.8%), followed by patients with gastroenterological (10.1%) and gynaecological malignancies (10.1%). Among patients with haematological malignancies, 75.6% of patients had chronic lymphocytic leukaemia or lymphomas and 19.5% patients had myeloid malignancies. On the basis of cancer and treatment modalities, we defined 4 cohorts: (i) patients with solid tumours (ST) receiving only chemotherapy (n = 63); (ii) ST patients receiving immunotherapy with or without chemotherapy (n = 16); (iii) ST patients receiving targeted or hormonal therapy (n = 79); and (iv) a combined group of haematological malignancy patients (n = 41) receiving either rituximab (n = 29), targeted therapy (n = 1), or an allogenic haematopoietic stem cell transplantation more than one year ago (n = 11).

### Sample collection and processing

Plasma samples were taken at the day of study inclusion (day 0, just before administration of the primer dose), at day 1 (the day after the primer dose), day 21 (just before administration of the booster dose), and day 28 (7 days after the booster dose). Serum samples were collected at day 49 (**Figure 1**). For detailed methods, refer to **Supplementary Information**.

**Figure 1.**
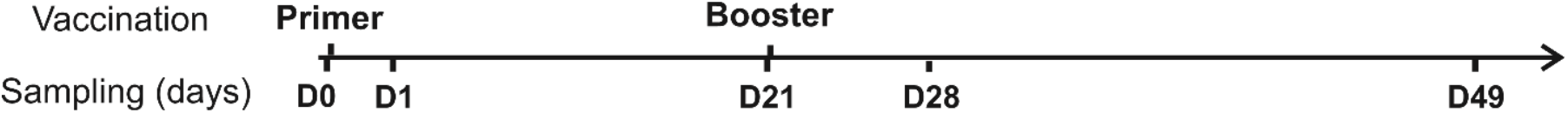
Timeline of the study. The BNT162b2 vaccine was administered on day 0 and 21. Heparin plasma samples for CCG analysis were collected on day 0 just prior to primer dose administration (D0), day 1 (D1), day 21 just prior to booster dose administration (D21), and day 28 (D28). For anti-RBD and anti-S1 serology, serum samples were collected on day 49 (D49) after the administration of the primer vaccine dose.

### Cytokine, chemokine and growth factor (CCG) measurements in plasma

CCGs were measured in plasma samples on a multiplex platform (Meso Scale Discovery (MSD), MD, USA) using off the shelf (V-plex) and customized (U-plex) panels, according to the manufacturer’s instructions, as previously described[19]. For detailed methods, refer to Supplementary Information.

In total, 36 CCGs relevant for SARS-CoV-2 infection or tumour growth and progression were measured. These constituted BDNF, bFGF, CRP, CTACK, FlT-1, IFN-β, IFN-γ, IL-1β, IL-1Ra, IL-2, IL-4, IL-5, IL-6, IL-8, IL-10, IL-13, IL-15, IL-16, IL-17A, IL-18, IL-21, IL-33, IP-10, monocyte chemoattractant protein (MCP)-1, PlGF, SAA, sICAM-1, VCAM-1, active and total (acid activated) tumour growth factor β (TGF-β), Tie-2, TNF-α, TSLP, VEGF-A, VEGF-C, and VEGF-D. Additionally, 5 extra CCGs were measured in a random subset of plasma samples collected from 100 cancer patients: G-CSF, granulocyte-macrophage colony-stimulating factor (GM-CSF), IL-7, IL-9, and MIP-1α.

### Anti-RBD IgG measurements in serum

Assessment of immunoglobulin G (IgG) antibody levels to the SARS-CoV-2 receptor-binding domain (RBD) antigen in this cohort has been previously described [5]. Anti-RBD antibody levels were measured in serum samples with an enzyme-linked immunosorbent assay (ELISA). A threshold for anti-RBD IgG of 200 IU/mL predicted a neutralization response required for 50% protection against symptomatic SARS-CoV-2 infection (99%-100% specificity at a sensitivity of 94.94%). As such, this threshold was used to differentiate high from low anti-SARS-CoV-2 serological responders, as described [5].

### Statistics

Group differences in CCG profiles of patients belonging to different treatment cohorts were investigated by Partial Least-Squares Discriminant Analysis (PLS-DA) using MetaboAnalyst (version 5.0). For this, data was primarily normalised with autoscaling and log10 transformation as described [19]. Different timepoints (before and after vaccination) were compared using a paired t-test on log-transformed data (SPSS v27). Low vs. high responders and patients with vs. without severe adverse events were compared using a two-sample t-test on log-transformed data (SPSS v27). To evaluate correlation between quantitative IgG levels and CCG concentrations, a Spearman correlation coefficient was utilized (R, version 4.1.0, http://www.rstudio.com/). A *p*-value <0.05 (uncorrected) was considered statistically significant.

For the identification of the main predictors of qualitative response (low/high responder), receiver operating characteristic (ROC) curves were constructed utilising MetaboAnalyst. To further predict low/high responder with a combined model of CCG levels, machine-learning-based Random Forest classifiers (RFC) were built (Python, package sklearn v2.0, ttp://scikit-learn.sourceforge.net). The main outcome variable was the development of an adequate immune response. To account for imbalanced groups, the Synthetic Minority Oversampling Technique (SMOTE, Python package imblearn 0.8.0) was utilized where 80% of the data was utilised as training set and the remaining 20% as test set. The models were bootstrapped 10 times and features for each model were selected based on 1) feature importance, 2) statistics from low vs. high responder, 3) Individual ROC curve analysis, and, 4) a Pearson correlation matrix for independence of variables. Confusion matrices and ROC curves were drawn to calculate area under the curve (AUROC) value to verify reliability and to evaluate the performance of the constructed models.

## Results

### Activation of early immune responses by BNT162b2 in cancer patients

We studied 41 key mediators of innate and/or adaptive immune responses as well as select growth factors relevant to immunity and/or cancer (see **Methods**). From this panel of CCGs, we observe a significant alteration of 23 CCGs after administration of the primer and/or the booster dose of the BNT162b2 vaccine in cancer patients under active treatment (Figure 1). Specifically, a day after the administration of the primer dose (day 1 vs. baseline day 0), anti-viral interferon (IFN) response molecules, such as IFN-γ induced protein 10 (IP-10/CXCL10), IFN-γ, and the T cell growth factor IL-9, were significantly upregulated (**Figure 2A, left panel**), suggesting, as expected, the importance of these immune mediators in the initial immune response to the vaccine.

**Figure 2.**
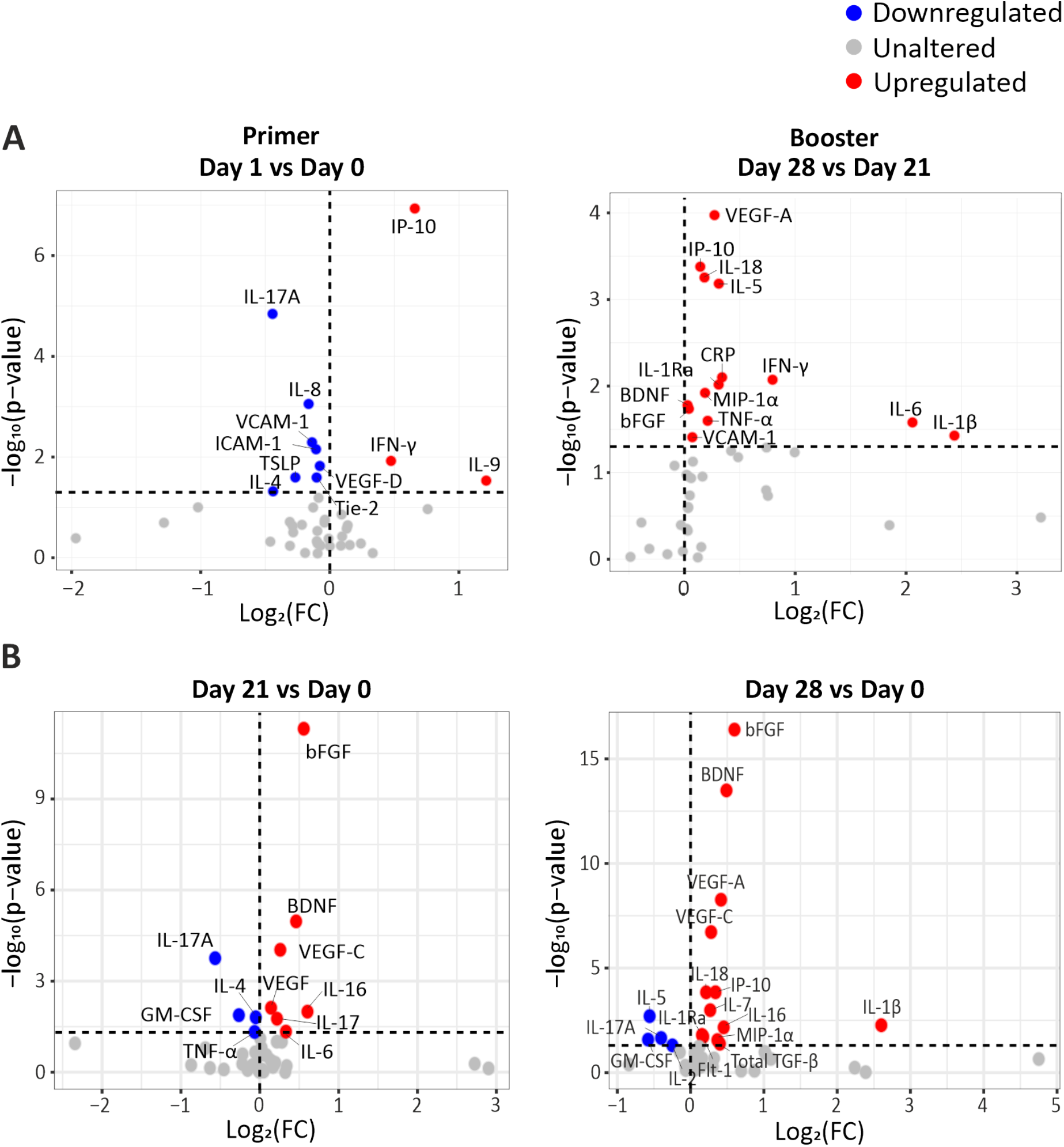
Volcano plots depicting differentially expressed CCGs **(A)** after the administration of the primer and booster doses compared to the CCG levels prior to vaccine administration and **(B)** comparing day 21 and day 28 with baseline day 0. *P*-values were calculated using paired t-test. The vertical dotted line represents no change. The horizontal dotted line represents a *p*-value of 0.05.

Importantly, the most prominently upregulated cytokines observed in our study, IP-10 and IFN-γ, were also observed to be prominently upregulated after the primer dose in a recent study on healthy volunteers [12]. On the other hand, we observe a downregulation of several CCGs after administration of the primer dose, such as IL-17A, IL-8, IL-4, thymic stromal lymphopoietin (TSLP), soluble vascular cell adhesion molecule 1 (VCAM-1), soluble intercellular adhesion molecule 1 (ICAM-1), Tie-2, and VEGF-D. Most of these analytes are crucially involved in the adaptive immune response or in cancer progression [25, 26]. For example, downregulation of TSLP, that has an important role in the maturation of T cell populations and enhancing Th2 responses [27], and of IL-4, a key Th2 cytokine with profound effects on B cell function, could be detrimental to the development of an adaptive immune response in the studied cancer patients (Figure 2A, left panel).

Seven days after booster dose administration, 14 CCGs were significantly elevated compared to the levels measured just before the booster dose administration. Interestingly, similarly to alterations observed after administration of the primer dose, upregulated CCGs included molecules responsible for the anti-viral IFN responses (IP-10, IFN-γ, and IFN-γ-inducing factor (IL-18)), but also inflammatory marker C-reactive protein (CRP), Th1 cytokines (tumour necrosis factor α (TNF-α), IL-1β, IL-6, and IL-1Ra), eosinophil promoting IL-5, and the chemokine macrophage inflammatory protein (MIP-1α), indicating activation of a wide range of immune responses despite the immunocompromised status of these patients. Upregulation of IP-10, IFN-γ, TNF-α, IL-6, IL-1Ra, CRP, and MIP-1α upon the administration of the booster dose has also been reported in healthy volunteers[12], although the upregulation in healthy volunteers is substantially higher (up to 20-fold) than those reported here in cancer patients (up to 2-fold). Notably, within the power of our study, none of the studied immunomodulatory and Treg CCGs (i.e., IL-10, IL-2, and IL-2Rα) were altered. On the other hand, levels of vascular injury marker VCAM-1 and angiogenesis markers brain-derived neurotrophic factor (BDNF), basic fibroblast growth factor (bFGF), and VEGF-A were upregulated 7 days after booster dose administration (day 28), compared to the levels studied at day 21 just before administration of the booster dose (**Figure 2A, right panel**).

We previously reported in this cohort that local or systemic adverse events (AEs) were mostly mild to moderate with only 3% (n = 5) and 6% (n =12) of patients experiencing severe local or systemic AEs after primer and booster dose, respectively[5]. Local reactogenicity was graded as mild, moderate, or severe. Systemic AEs were recorded according to the Common Terminology Criteria for Adverse Events version 5.0 (CTCAE v5.0; graded 0–5; 5 being death). Additionally, we investigated whether CCG responses are different in patients who developed severe AE: only placental growth factor (PlGF) was observed to be significantly downregulated after the primer dose in an uncorrected paired t-test statistics (*p* = 0.027) and was not significant after post-hoc false discovery rate correction. These data fit well with studies suggesting that systemic adverse events noted after vaccination in cancer patients are not necessarily vaccine related [5, 28].

Lastly, as several angiogenic markers are important factors in tumour progression, we studied their differences between day 1 and day 21 (pre-booster vaccination) and between day 1 and day 28. Angiogenic markers bFGF, BDNF, and VEGF-A were significantly increased in both comparisons (**Figure 2B**). An independent regression analysis also showed that these angiogenic markers along with VEGF receptor vascular endothelial growth factor receptor (Flt-1) were significantly increasing over time since the primer dose administration (**Supplementary Figure 1**). However, in the absence of a non-vaccinated cancer patient group it is difficult to ascertain whether this significant increase in angiogenic markers BDNF, bFGF, Flt-1, and VEGF-A is the effect of vaccination or part of natural progression of cancer in these patients.

### Type of cancer therapy does not majorly alter the CCG profile induced by the BNT162b2 mRNA vaccine

An inadequate IgG immune response to the BNT162b2 mRNA vaccine was reported especially in haematological malignancy patients and notably in those receiving rituximab, an anti-CD20 B cell blocker [5]. We thus first questioned whether CCG profiles could discriminate haematological cancer patients receiving rituximab from haematological malignancy patients receiving stem cell transplantation, the other major treatment modality for haematological malignancy patients studied in this report, or from all other cancer and treatment groups combined. A significant discrimination was observed at day 1 for haematological cancer patients with or without rituximab (accuracy = 87%; R^2^ = 0.67; Q^2^ = 0.30), but was not observed at other timepoints, nor was observed at any timepoint when combining the groups of solid cancer and non-rituximab-treated haematological cancer patients (**Supplementary Figure 2**). As other treatment modalities, especially chemotherapy for patients with solid tumours, also showed a diminished immune response, we performed a similar discriminant analysis that showed no significant underlying difference in CCG profiles at any timepoint (**Figure 3; Supplementary Figure 3**).

**Figure 3.**
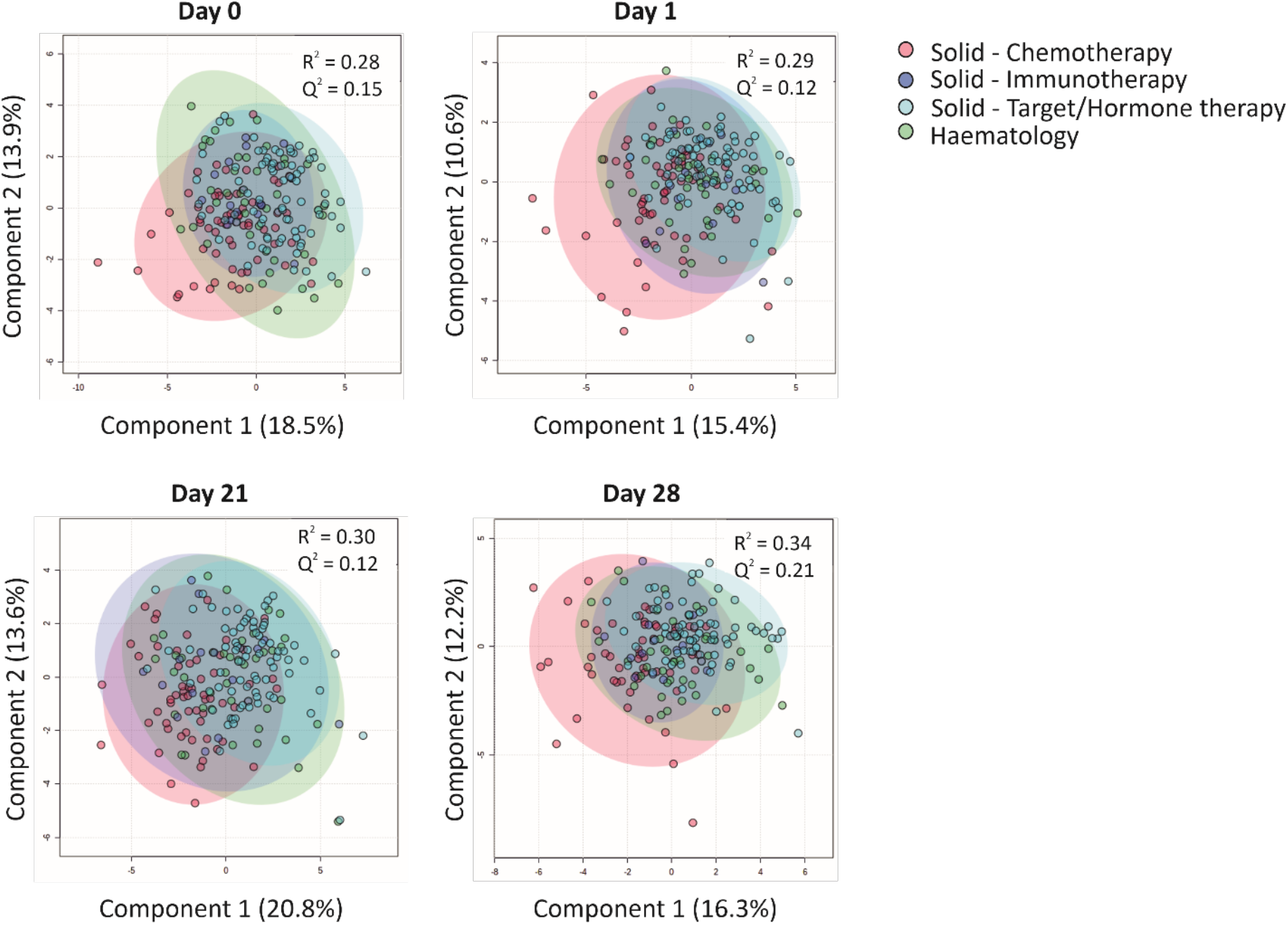
CCG analysis in patients undergoing different treatment regimens. Cluster analyses of CCGs at different timepoints with a Partial Least Squares-Discriminant Analysis (PLS-DA) reveal minor differences between patients undergoing distinct types of anti-cancer therapies. Haematological patients included patients receiving rituximab or patients who received an allogeneic hematopoietic stem cell transplantation at least 1 year before the primer dose vaccination.

Studying individual cytokines in a difference of mean analysis revealed that the only cytokine linked to vaccine administration and upregulated in all treatment groups was IP-10, which is indicative of an effective anti-viral immune response. Moreover, IP-10-regulator IFN-γ was also upregulated in patients with solid tumours treated with targeted or hormonal therapy (Supplementary Figure 3). Surprisingly, neutrophil chemoattractant IL-8 was downregulated in both haematological malignancy patients and patients with solid tumours treated with targeted/hormonal therapy. Moreover, all groups of patients with solid tumours demonstrated a significant downregulation of IL-17A, a pro-inflammatory cytokine involved mainly in the activation of neutrophils. Lastly, the solid tumour cohort treated with chemotherapy or targeted/hormone therapy showed a significant increase of VEGF-C, bFGF, and BDNF over 21 days (Supplementary Figure 3). These data indicate that except for rituximab-treated haematological malignancy groups that behave differently at day 1, type of cancer therapy is not a major driver for the observed CCG profiles induced by the BNT162b2 vaccination.

### CRP, IL-15, IL-18, and PlGF predict a low BNT162b2 immune response in cancer patients

Due to the limited ability of some patients with solid or haematological malignancies to develop a protective antibody response, we aimed to identify a unique CCG signature that could differentiate good from poor responders to BNT162b2 vaccination in cancer patients. For this, we examined the relationship between alterations in the studied CCGs at all sampling timepoints (day 0, day 1, day 21, and day 28) with levels of anti-RBD titres measured 28 days after the administration of the booster dose of the BNT162b2 vaccine (day 49) (Figure 1). This was done following several approaches. First, we utilized anti-RBD titres measured at day 49 as a continuous variable and correlated with CCGs at all studied timepoints. Amongst others, BDNF, VEGF-C, IFN-γ, IFN-β, and ICAM-1 were significantly positively associated with anti-RBD titres at one or several timepoints (**Figure 4A**). Additionally, bFGF, PIGF, IL-18, granulocyte colony stimulating factor (G-CSF), and pro-inflammatory cytokines IL-15 and IL-16 were significantly negatively associated with anti-RBD titres (Figure 4A). These data suggest that pre-existing and sustained CCG signatures in patients with solid and haematological malignancies can be predictive of the quantitative antibody response post BNT162b2 vaccination.

**Figure 4.**
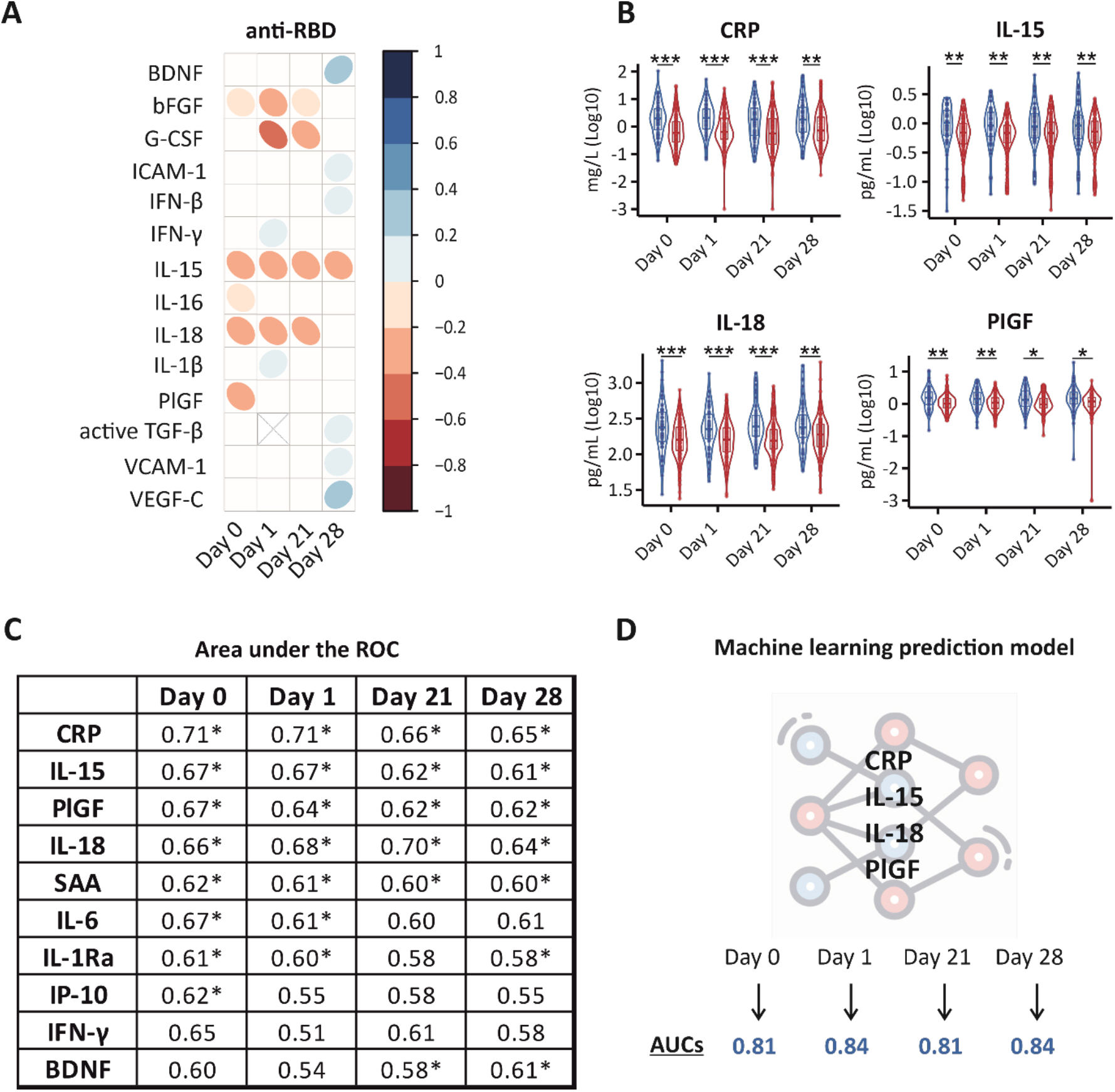
Prediction models for poor BNT162b2 immune response in cancer patients. **(A)** Correlation matrix depicting the correlation between CCG measurements (log10 transformed) and quantitative anti-RBD and anti-S1 IgG measurements at day 49. IgG antibody levels to SARS-CoV-2 RBD antigen were assessed with an enzyme-linked immunosorbent assay for quantitative detection of IgG antibody levels to SARS-CoV-2 receptor-binding domain (RBD) antigen. Only CCGs with significant correlations are shown. **(B)** Violin plots for CCGs significantly different between low (< 200 IU/mL) (blue) and high (≥ 200 IU/mL) (red) responders to the BNT162b2 vaccine. A low/high responder threshold (anti-RBD IgG titre of 200 IU/mL) used in this study predicts a neutralization response required for 50% protection against symptomatic SARS-CoV-2 infection (99%-100% specificity at a sensitivity of 95%). * *p* < 0.05, ** *p* < 0.01, *** *p* < 0.001. **(C)** Area Under the Receiver Operating Characteristic (AUROC) values for 10 predictors of the binary IgG response as low or high responders at day 0, day 1, day 21, and day 28. * Denotes significant *p*-values of at least < 0.05. **(D)** Random Forest Classifier predicted a model where a combination of CRP, IL-15, IL-18, and PlGF levels measured at right before vaccine administration (day 0) and at day 1, day 21, and day 28 after the primer dose predicted low responders and high responders with high accuracy (AUCs depicts averages of 10 individually constructed ROCs).

Since the clinical question addressed the level of protection conferred by vaccination, we further utilised a threshold of 200 IU/mL shown to predict a neutralisation response conferring 50% protection against SARS-CoV-2 infection [5]. Examining the ability of CCGs to predict low responders (< 200 IU/mL) from high responders (≥ 200 IU/mL), 4 CCGs were identified to be significantly different at all studied timepoints that included CRP, IL-15, IL-18, and PlGF (**Figure 4B**).

Further, area under the curve receiver operating characteristic (AUROC) analysis was performed to discriminate between low and high responders. AUROC was constructed for each CCG and the top discriminant CCGs were utilised to build models. Performance was studied for each timepoint to assess for the capability of the model to sustain at all studied timepoints. Prior to vaccine administration, the upregulated inflammatory marker CRP showed the highest predictive value for poor response to the vaccine administration followed by NK-cell inducer IL-15, PlGF, IL-6, IL-18, and serum amyloid A (SAA). One day after administration of the primer dose, CRP, IL-18, IL-15, PlGF, IL6, and SAA remained in the signature predicting a worse qualitative antibody response. Prior to administration of the booster dose, the signature included CRP, IL-15, IL-18, PlGF, and SAA (**Figure 4C**). CRP on its own was not a good classifier with a cut-off of 1 mg/L leading to a sensitivity and specificity of 72% and 61%, respectively (see **Supplementary Table 2** for more details). Lastly, we performed Random Forest classification that validated the signature consisting of CRP, IL-15, IL-18, and PlGF, differentiating low from high anti-SARS-CoV-2 BNT162b2 vaccine responders with more than 80% accuracy. Interestingly, this signature was maintained until day 28 after the administration of the primer dose (**Figure 4D, Supplementary Figure 4**).

## Discussion

In this study, we show an alteration of a diverse group of inflammatory mediators and growth factors that includes interferons, Th1, Th2, and Th17 cytokines, as well as some markers of angiogenesis and vascular injury in a heterogeneous population of patients with solid or haematological malignancies vaccinated with BNT162b2. Some of the CCGs, including IP-10, IFN-γ, TNF-α, IL-6, IL-1Ra, CRP, MIP-1α, and VEGF-A have previously been shown to be upregulated upon vaccine administration in healthy individuals, assessed on the same immunoassay platform as employed in this study [12]. Additionally, an increase in IFN-γ and IP-10 levels was also observed in an elderly population upon the administration of the BNT162b2 vaccine but to a larger magnitude to that reported in our cohort [23]. These data suggest that BNT162b2 vaccine administration in cancer patients can generally elicit an anti-SARS-CoV-2-driven immune response that is similar in pattern, but not in magnitude, compared to healthy individuals.

Notably, we also identified CCG alterations unique to either solid (i.e., CRP, SAA, IL-1Ra, IL-17A, IL-4, IL-5, IL-9, and MIP-1α) or haematological (i.e., TSLP, IL-8, sICAM-1, and VCAM-1) malignancies as a response to COVID-19 vaccine. Several of these cytokines are known to have a significant tumour-promoting effect, implying that vaccine administration might temporarily favour tumour progression. However, due to the national legislation on vaccination of cancer patients that did not allow a matched group of non-vaccinated cancer patients, it is difficult to ascertain whether this significant increase in cancer-promoting factors is the effect of vaccination or part of natural cancer progression in these patients.

All cohorts of patients with solid and haematological malignancies undergoing different treatment regimens developed anti-viral interferon responses after vaccination with BNT162b2. However, with exception of the rituximab treatment cohort, no major underlying differences in CCG profiles were identified between different cancer or treatment groups at any timepoint. These data suggest that despite having different tumour types and undergoing different therapies, patients respond similarly to vaccination with BNT162b2.

Previous studies have shown that patients with certain cancers, including but not limited to, advanced cancers and B cell haematological malignancies, develop low or even absent antibody titres not only after SARS-CoV-2 infection, but also after SARS-CoV-2 vaccination [5, 7-11, 29-32]. In line with the major aim of this study, we identified a unique immune signature based on upregulated CRP, IL-15, IL-18, and PlGF can be used to identify patients who did not sufficiently respond to vaccination with BNT162b2 vaccine. The signature was present at different studied timepoints before or after vaccination and was not majorly affected by different anti-cancer treatments. We believe that this unique biomarker would not only be useful for clinicians in identifying cancer patients at increased risk of developing SARS-CoV-2 for better patient care, but also be able to guide health policies in categorising cancer patients in need of enhancer vaccine doses or pre-exposure prophylaxis with synthetic monoclonal antibodies to protect potential non-responders to the BNT162b2 vaccine.

Lastly, our data also suggest that pro-inflammatory cytokines and growth factors interact to dictate an inherent immune response in cancer patients that could generally render them refractive to other immune interventions. Whether the identified signature or similar immune-based CCG profile can be predictive of primary resistance to immunotherapy, observed in approximately 12% of the patients [33], remains open to future investigations.

## Data Availability

All data produced in the present study are available upon reasonable request to the authors.

## Author contributions

Conceptualization: SKS, PvD, MP; Funding acquisition: SKS, PvD, MP, ET; Overall study supervision: SKS; Clinical data and sample collection: LV, LAT, GV, SR, IVdM, SDK, YD, GM, BP; Biobanking: MH; CCG analysis: AK, FDW, AG, AH; Serology and seroneutralisation analyses: PP, KN, KA, MVDB, ID, MG; Statistical analysis: AK, FDW, AG, AH; Data curation: AK, AG, ER; Data interpretation: SKS, PvD, MP, TV, SMK, AK, FDW, AG, AH, MB; Manuscript writing: SKS, AK, FDW, AG, AH, MB; All authors read, gave input, and approved the final manuscript.

## References

1. Goldberg Y, Mandel M, Bar-On YM, Bodenheimer O, Freedman L, Haas EJ, et al. Waning Immunity after the BNT162b2 Vaccine in Israel. N Engl J Med. 2021;385(24):e85. https://doi.org:10.1056/NEJMoa2114228

2. Polack FP, Thomas SJ, Kitchin N, Absalon J, Gurtman A, Lockhart S, et al. Safety and Efficacy of the BNT162b2 mRNA Covid-19 Vaccine. N Engl J Med. 2020;383(27):2603–15. https://doi.org:10.1056/NEJMoa2034577

3. Thomas SJ, Moreira ED, Jr., Kitchin N, Absalon J, Gurtman A, Lockhart S, et al. Safety and Efficacy of the BNT162b2 mRNA Covid-19 Vaccine through 6 Months. N Engl J Med. 2021;385(19):1761–73. https://doi.org:10.1056/NEJMoa2110345

4. Teijaro JR, Farber DL. COVID-19 vaccines: modes of immune activation and future challenges. Nature reviews Immunology. 2021;21(4):195–7. https://doi.org:10.1038/s41577-021-00526-x

5. Peeters M, Verbruggen L, Teuwen L, Vanhoutte G, Vande Kerckhove S, Peeters B, et al. Reduced humoral immune response after BNT162b2 coronavirus disease 2019 messenger RNA vaccination in cancer patients under antineoplastic treatment. ESMO Open. 2021;6(5):100274. https://doi.org:10.1016/j.esmoop.2021.100274

6. Cortés A, Casado JL, Longo F, Serrano JJ, Saavedra C, Velasco H, et al. Limited T cell response to SARS-CoV-2 mRNA vaccine among patients with cancer receiving different cancer treatments. Eur J Cancer. 2022;166:229–39. https://doi.org:10.1016/j.ejca.2022.02.017

7. Massarweh A, Eliakim-Raz N, Stemmer A, Levy-Barda A, Yust-Katz S, Zer A, et al. Evaluation of Seropositivity Following BNT162b2 Messenger RNA Vaccination for SARS-CoV-2 in Patients Undergoing Treatment for Cancer. JAMA Oncol. 2021;7(8):1133–40. https://doi.org:10.1001/jamaoncol.2021.2155

8. Maneikis K, Šablauskas K, Ringelevičiūtė U, Vaitekėnaitė V, Čekauskienė R, Kryžauskaitė L, et al. Immunogenicity of the BNT162b2 COVID-19 mRNA vaccine and early clinical outcomes in patients with haematological malignancies in Lithuania: a national prospective cohort study. The Lancet Haematology. 2021;8(8):e583–e92. https://doi.org:10.1016/s2352-3026(21)00169-1

9. Fendler A, Shepherd STC, Au L, Wilkinson KA, Wu M, Byrne F, et al. Adaptive immunity and neutralizing antibodies against SARS-CoV-2 variants of concern following vaccination in patients with cancer: the CAPTURE study. Nature Cancer. 2021;2(12):1305–20. https://doi.org:10.1038/s43018-021-00274-w

10. Linardou H, Spanakis N, Koliou GA, Christopoulou A, Karageorgopoulou S, Alevra N, et al. Responses to SARS-CoV-2 Vaccination in Patients with Cancer (ReCOVer Study): A Prospective Cohort Study of the Hellenic Cooperative Oncology Group. Cancers (Basel). 2021;13(18). https://doi.org:10.3390/cancers13184621

11. Chung DJ, Shah GL, Devlin SM, Ramanathan LV, Doddi S, Pessin MS, et al. Disease- and Therapy-Specific Impact on Humoral Immune Responses to COVID-19 Vaccination in Hematologic Malignancies. Blood Cancer Discov. 2021;2(6):568–76. https://doi.org:10.1158/2643-3230.Bcd-21-0139

12. Bergamaschi C, Terpos E, Rosati M, Angel M, Bear J, Stellas D, et al. Systemic IL-15, IFN-gamma, and IP-10/CXCL10 signature associated with effective immune response to SARS-CoV-2 in BNT162b2 mRNA vaccine recipients. Cell reports. 2021;36(6):109504. https://doi.org:10.1016/j.celrep.2021.109504

13. Miyashita Y, Yoshida T, Takagi Y, Tsukamoto H, Takashima K, Kouwaki T, et al. Circulating extracellular vesicle microRNAs associated with adverse reactions, proinflammatory cytokine, and antibody production after COVID-19 vaccination. NPJ Vaccines. 2022;7(1):16. https://doi.org:10.1038/s41541-022-00439-3

14. Monin MB, Marx B, Meffert L, Boesecke C, Rockstroh JK, Strassburg CP, et al. SARS-CoV-2 PrEP complicates antibody testing after vaccination: a call for awareness. Ann Hematol. 2022;101(9):2089–90. https://doi.org:10.1007/s00277-022-04859-y

15. Pagano L, Salmanton-García J, Marchesi F, Blennow O, Gomes da Silva M, Glenthøj A, et al. Breakthrough COVID-19 in vaccinated patients with hematologic malignancies: results from EPICOVIDEHA survey. Blood. 2022. https://doi.org:10.1182/blood.2022017257

16. Grivennikov SI, Greten FR, Karin M. Immunity, Inflammation, and Cancer. Cell. 2010;140(6):883–99. https://doi.org/10.1016/j.cell.2010.01.025

17. Mantovani A, Allavena P, Sica A, Balkwill F. Cancer-related inflammation. Nature. 2008;454(7203):436–44. https://doi.org:10.1038/nature07205

18. Lan T, Chen L, Wei X. Inflammatory Cytokines in Cancer: Comprehensive Understanding and Clinical Progress in Gene Therapy. Cells. 2021;10(1). https://doi.org:10.3390/cells10010100

19. De Winter Fhr, Hotterbeekx A, Huizing MT, Konnova A, Fransen E, Jongers B, et al. Blood Cytokine Analysis Suggests That SARS-CoV-2 Infection Results in a Sustained Tumour Promoting Environment in Cancer Patients. Cancers (Basel). 2021;13(22). https://doi.org:10.3390/cancers13225718

20. Skibinski DAG, Jones LA, Zhu YO, Xue LW, Au B, Lee B, et al. Induction of Human T-cell and Cytokine Responses Following Vaccination with a Novel Influenza Vaccine. Scientific reports. 2018;8(1):18007. https://doi.org:10.1038/s41598-018-36703-7

21. Williams CM, Roy S, Califano D, McKenzie ANJ, Metzger DW, Furuya Y. The Interleukin-33-Group 2 Innate Lymphoid Cell Axis Represents a Potential Adjuvant Target To Increase the Cross-Protective Efficacy of Influenza Vaccine. Journal of virology. 2021;95(22):e0059821. https://doi.org:10.1128/jvi.00598-21

22. Tahtinen S, Tong AJ, Himmels P, Oh J, Paler-Martinez A, Kim L, et al. IL-1 and IL-1ra are key regulators of the inflammatory response to RNA vaccines. Nature immunology. 2022;23(4):532–42. https://doi.org:10.1038/s41590-022-01160-y

23. Lee HK, Knabl L, Knabl L, Kapferer S, Pateter B, Walter M, et al. Robust immune response to the BNT162b mRNA vaccine in an elderly population vaccinated 15 months after recovery from COVID-19. medRxiv. 2021. https://doi.org:10.1101/2021.09.08.21263284

24. Funakoshi Y, Yakushijin K, Ohji G, Hojo W, Sakai H, Takai R, et al. Safety and immunogenicity of the COVID-19 vaccine BNT162b2 in patients undergoing chemotherapy for solid cancer. J Infect Chemother. 2022;28(4):516–20. https://doi.org:10.1016/j.jiac.2021.12.021

25. Kong D-H, Kim YK, Kim MR, Jang JH, Lee S. Emerging Roles of Vascular Cell Adhesion Molecule-1 (VCAM-1) in Immunological Disorders and Cancer. Int J Mol Sci. 2018;19(4):1057.

26. Landskron G, De la Fuente M, Thuwajit P, Thuwajit C, Hermoso MA. Chronic Inflammation and Cytokines in the Tumor Microenvironment. Journal of immunology research. 2014;2014:149185. https://doi.org:10.1155/2014/149185

27. Kitajima M, Lee HC, Nakayama T, Ziegler SF. TSLP enhances the function of helper type 2 cells. European journal of immunology. 2011;41(7):1862–71. https://doi.org:10.1002/eji.201041195

28. Walle T, Bajaj S, Kraske JA, Rösner T, Cussigh CS, Kälber KA, et al. Cytokine release syndrome-like serum responses after COVID-19 vaccination are frequent and clinically inapparent under cancer immunotherapy. Nat Cancer. 2022. https://doi.org:10.1038/s43018-022-00398-7

29. van Dam PA, Huizing M, Papadimitriou K, Prenen H, Peeters M. High mortality of cancer patients in times of SARS-CoV-2: Do not generalize! Eur J Cancer. 2020;138:225–7. https://doi.org:10.1016/j.ejca.2020.07.021

30. van Dam P, Huizing M, Roelant E, Hotterbeekx A, De Winter Fhr, Kumar-Singh S, et al. Immunoglobin G/total antibody testing for SARS-CoV-2: A prospective cohort study of ambulatory patients and health care workers in two Belgian oncology units comparing three commercial tests. Eur J Cancer. 2021;148:328–39. https://doi.org:10.1016/j.ejca.2021.02.024

31. Fendler A, Au L, Shepherd STC, Byrne F, Cerrone M, Boos LA, et al. Functional antibody and T cell immunity following SARS-CoV-2 infection, including by variants of concern, in patients with cancer: the CAPTURE study. Nat Cancer. 2021;2(12):1321–37. https://doi.org:10.1038/s43018-021-00275-9

32. Overheu O, Quast DR, Schmidt WE, Sakinç-Güler T, Reinacher-Schick A. Low Serological Prevalence of SARS-CoV-2 Antibodies in Cancer Patients at a German University Oncology Center. Oncol Res Treat. 2022;45(3):112–7. https://doi.org:10.1159/000520572

33. Haslam A, Prasad V. Estimation of the Percentage of US Patients With Cancer Who Are Eligible for and Respond to Checkpoint Inhibitor Immunotherapy Drugs. JAMA Netw Open. 2019;2(5):e192535. https://doi.org:10.1001/jamanetworkopen.2019.2535

