## Supplementary methods, tables and figures for "Predictive model for BNT162b2 vaccine response in cancer patients based on cytokines and growth factors"

#### Sample collection and processing

Whole blood was prospectively collected in heparin blood and serum collection tubes. Within 3 hours of blood collection, plasma was prepared by centrifuging twice at 1900×g for 10 minutes without brakes. Blood in the serum collection tubes was allowed to clot thoroughly for 60 minutes and serum was prepared by centrifuging at 1300×g for 10 minutes without brakes. Aliquots were flash frozen in liquid nitrogen and stored in the Biobank of Antwerp University Hospital at -80°C until further analysis.

#### Cytokine, chemokine and growth factor (CCG) measurements in plasma

CCGs were measured in plasma samples on a multiplex platform (Meso Scale Discovery (MSD), MD, USA) using off the shelf (V-plex) and customized (U-plex) panels, following manufacturer instructions. Briefly, 96-well plates of the U-plex panels were coated with a capturing antibody coupled to a linker for one hour. The vascular injury panel (K15198D) was washed before use. The angiogenesis panel (K15190D) was first blocked with blocking buffer for one hour. Afterwards, all plates were washed three times with PBS-Tween (0.05%). Samples were incubated for one hour (except for the angiogenesis and the vascular injury panels, where two hours of incubation were performed), after which the plates were washed another three times. Detection antibody with a SULFO-TAG was added and after another one-hour incubation step (two hours for the angiogenesis panel), the plates were washed and read with MSD reading buffer on the QuickPlex SQ 120 (MSD) as previously described[18].

In total, 36 CCGs relevant for SARS-CoV-2 infection or tumour growth and progression were measured. These constituted BDNF, bFGF, CRP, CTACK, FlT-1, IFN-β, IFN-γ, IL-1β, IL-1Ra, IL-2, IL-4, IL-5, IL-6, IL-8, IL-10, IL-13, IL-15, IL-16, IL-17A, IL-18, IL-21, IL-33, IP-10, monocyte chemoattractant protein (MCP)-1, PlGF, SAA, sICAM-1, VCAM-1, active and total (acid activated) tumour growth factor β (TGF-β), Tie-2, TNF-α, TSLP, VEGF-A, VEGF-C, and VEGF-D. Additionally, 5 extra CCGs were measured in a random subset of plasma samples collected from 100 cancer patients: G-CSF, granulocyte-macrophage colony-stimulating factor (GM-CSF), IL-7, IL-9, and MIP-1α.

### SUPPLEMENTARY FIGURES

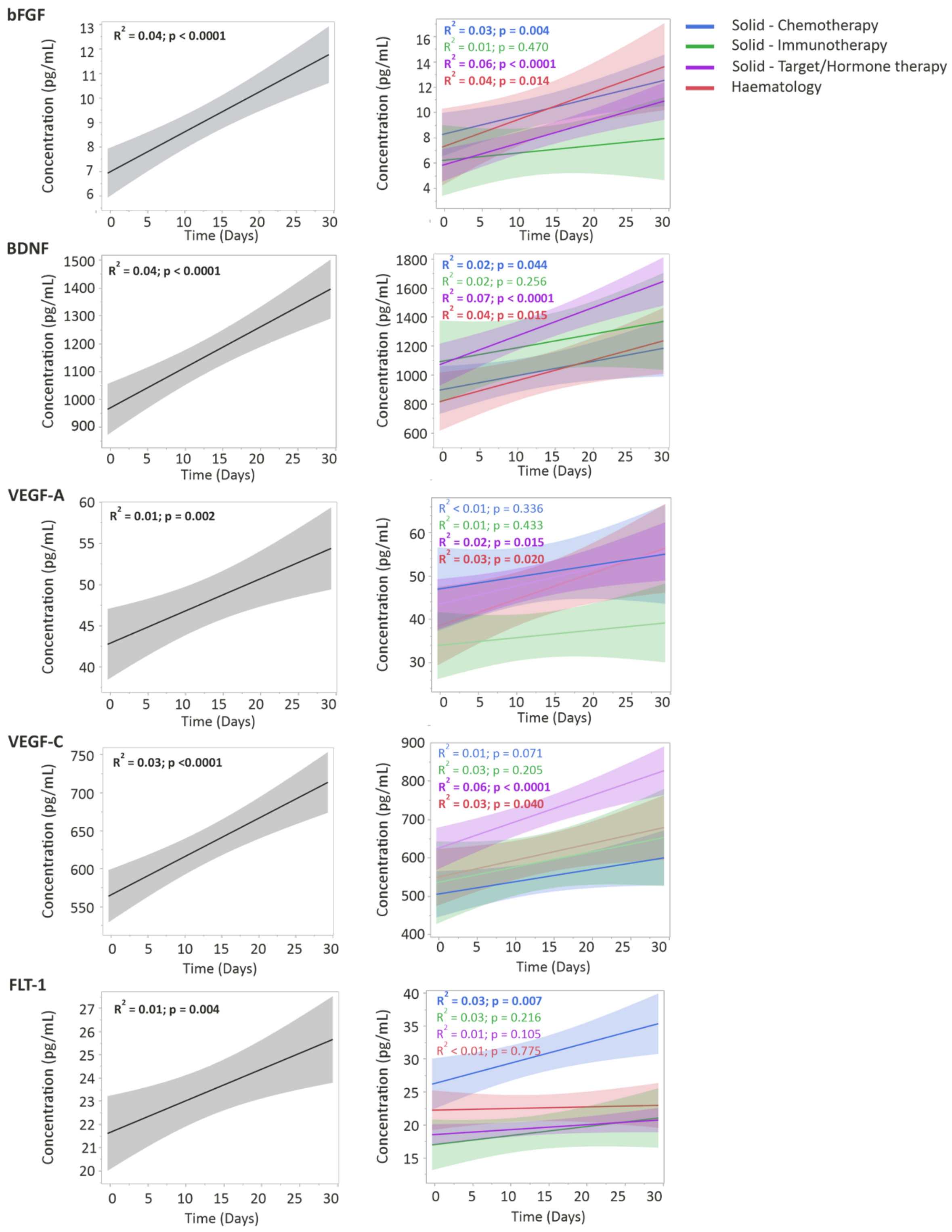


**Supplementary Figure 1.** Temporal evolution of cytokines, chemokines and growth factors (CCGs) altered in vaccinated cancer patients. Time is represented as days since primer dose vaccination. *P*-values in the graph refer to significance of the slope of the regression lines.

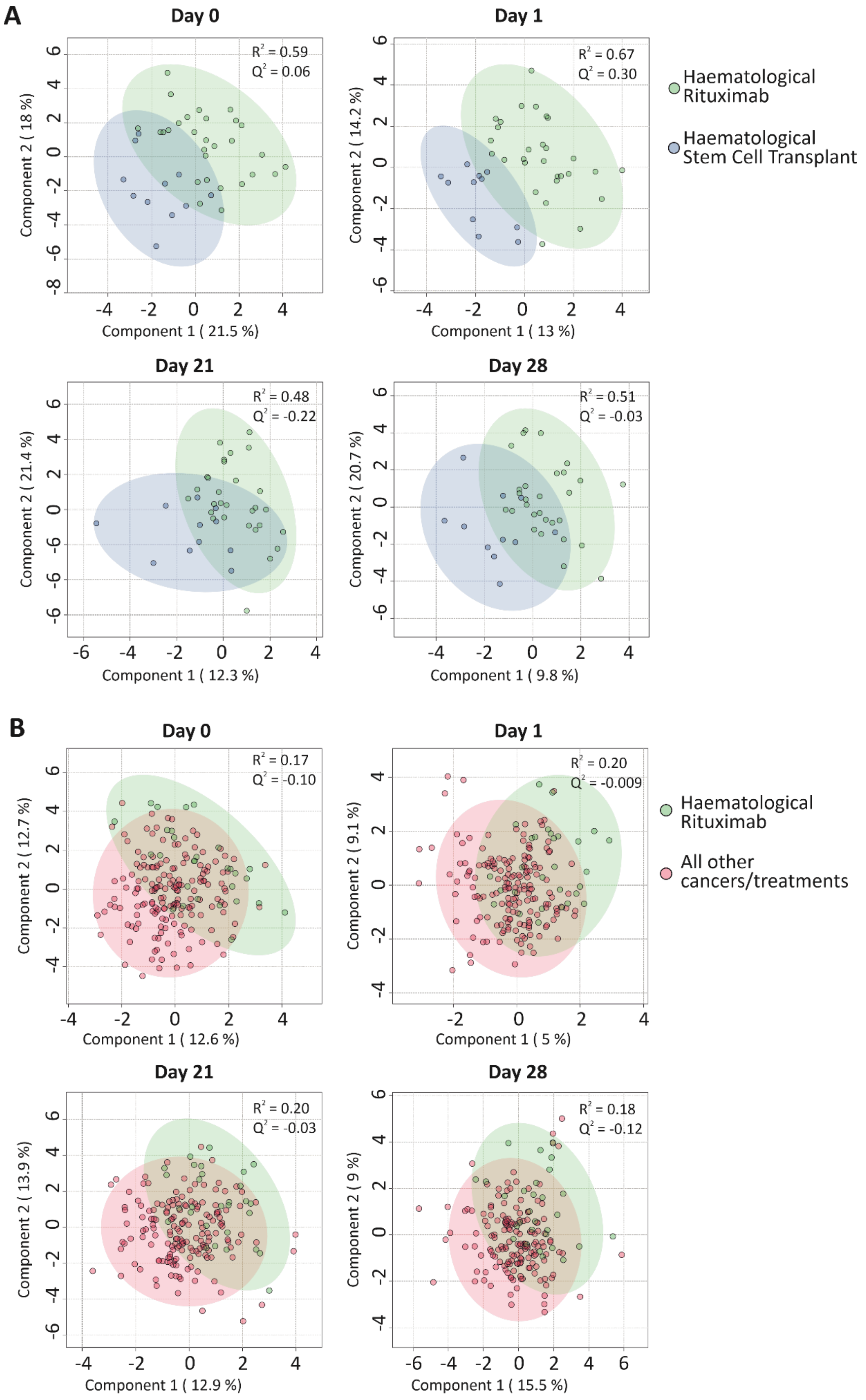


**Supplementary Figure 2**. CCG analysis in patients with and without haematological malignancies. Cluster analyses of CCGs at different timepoints with partial least squares-discriminant analysis (PLS-DA) reveal (A) differences between patients with haematological malignancies treated with rituximab and patients that received a stem cell transplantation, or (B) between patients with haematological malignancies treated with rituximab versus all other cancers/treatments group

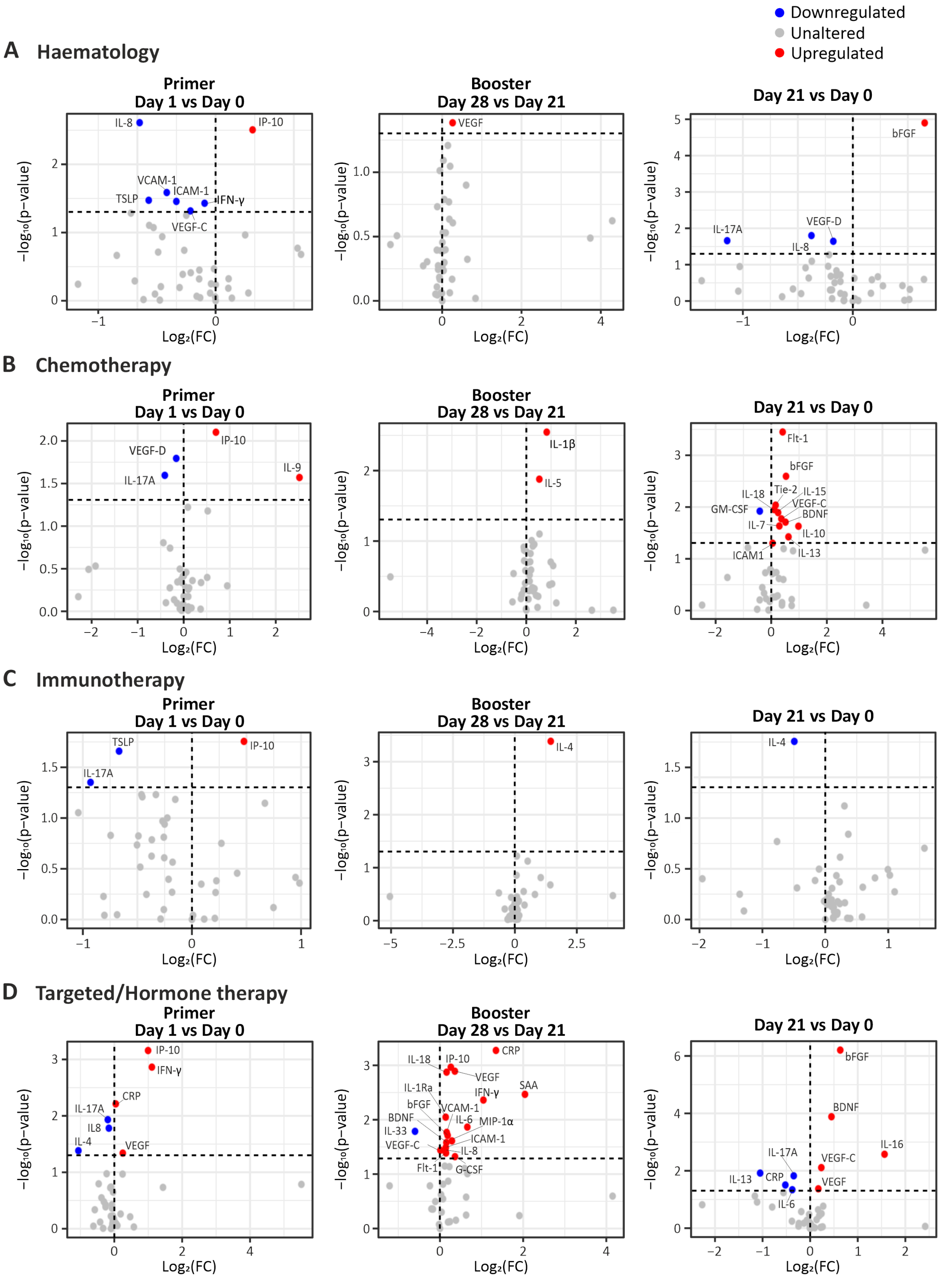


**Supplementary Figure 3**. Volcano plots depicting differentially expressed CCGs after the administration of the primer and booster doses compared to the CCG levels prior to vaccine administration (A) in cancer with haematological malignancies, (B) patients with solid cancers treated with chemotherapy, (C) immunotherapy and (D) targeted or hormonal therapy. *P*-values were calculated using paired t-test. The vertical dotted line represents no change. The horizontal dotted line represents a *p*-value of 0.05.

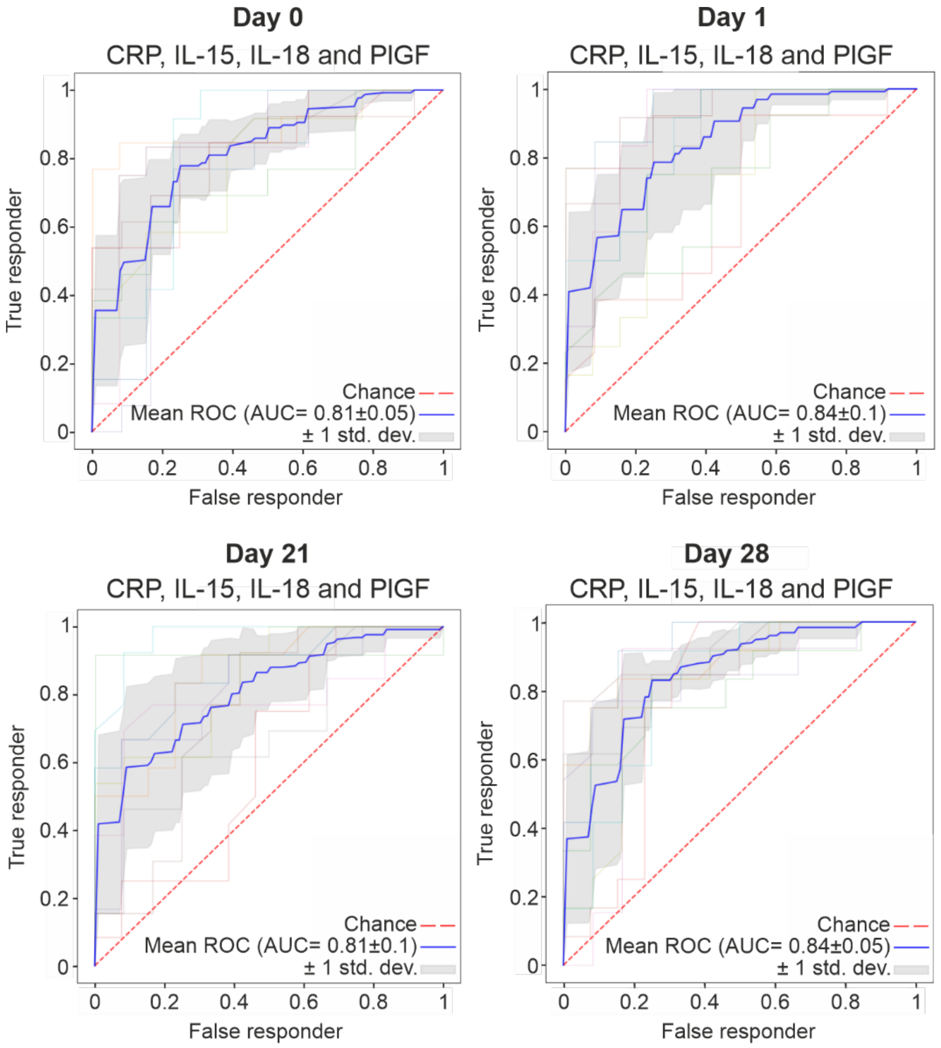


**Supplementary Figure 4**. ROC curves for the combination of CRP, IL-15, IL-18 and PlGF in a random forest classifier model with Synthetic Minority Oversampling Technique (SMOTE) for the prediction of the qualitative IgG response (responder or non-responder) are depicted for day 0, day 1, day 21 and day 28.

**Supplementary Table 1**. Patient characteristics (adapted from Peeters *et al.*) *Percentage of total patients with solid tumours.

| Demographics | Target/hormone therapy (*n* = 79) | Immuno-therapy (*n* = 16) | Chemo-therapy (n = 63) | Haema-tological (*n* = 41) | Overall (n = 199) |
|---|---|---|---|---|---|
| Sex, *n* (%) | | | | | |
| Female | 70 (88.6) | 4 (25.0) | 43 (68.3) | 17 (41.5) | 134 (67.3) |
| Male | 9 (11.4) | 12 (75.0) | 20 (31.7) | 24 (58.5) | 65 (32.7) |
| Age, years | | | | | |
| Mean (SD) | 59.5 (12.1) | 68.3 (8.09) | 60.0 (13.2) | 61.2 (11.5) | 60.7 (12.2) |
| Median (range) | 60.0 (31.0-86.0) | 69.5 (56.0-84.0) | 61.0 (26.0-88.0) | 63.0 (25.0-79.0) | 62.0 (25.0-88.0) |
| BMI | | | | | |
| Mean (SD) | 25.7 (4.74) | 27.0 (4.13) | 25.5 (5.19) | 25.2 (3.88) | 25.6 (4.67) |
| Median (range) | 25.5 (17.8-40.0) | 26.9 (19.7-34.5) | 24 (18.9-44.8) | 24.4 (17.1-35.5) | 25.1 (17.1-44.8) |
| Missing, *n* (%) | 0 (0) | 0 (0) | 3 (4.8) | 2 (4.9) | 5 (2.5) |
| ECOG score, *n* (%) | | | | | |
| 0 | 73 (92.4) | 11 (68.8) | 48 (76.2) | 38 (92.7) | 170 (85.4) |
| 1 | 6 (7.6) | 5 (31.2) | 13 (20.6) | 3 (7.3) | 27 (13.6) |
| 2 | 0 (0) | 0 (0) | 1 (1.6) | 0 (0) | 1 (0.5) |
| Missing | 0 (0) | 0 (0) | 1 (1.6) | 0 (0) | 1 (0.5) |
| Autoimmune disease, *n* (%) | 4 (5.1) | 0 (0) | 1 (1.6) | 3 (7.3) | 8 (4.0) |
| Kidney disease, *n* (%) | 1 (1.3) | 1 (6.2) | 5 (7.9) | 1 (2.4) | 8 (4.0) |
| Hypertension, *n* (%) | 20 (25.3) | 4 (25.0) | 22 (34.9) | 8 (19.5) | 54 (27.1) |
| Diabetes, *n* (%) | 3 (3.8) | 2 (12.5) | 10 (15.9) | 5 (12.2) | 20 (10.1) |
| Coronary disease, *n* (%) | 4 (5.1) | 2 (12.5) | 10 (15.9) | 7 (17.1) | 23 (11.6) |
| Smoking status, *n* (%) | | | | | |
| Current smoker | 5 (6.3) | 1 (6.2) | 5 (7.9) | 2 (4.9) | 13 (6.5) |
| Former smoker | 21 (26.6) | 11 (68.8) | 21 (33.3) | 18 (43.9) | 71 (35.7) |
| Non-smoker | 51 (64.6) | 3 (18.8) | 29 (46.0) | 21 (51.2) | 104 (52.3) |
| Missing | 3 (2.5) | 1 (6.2) | 8 (12.7) | 0 (0) | 11 (5.5) |
| Stage, *n* (%) | | | | | |
| I | 20 (25.3) | 0 (0) | 6 (9.5) | NA | 26 (16.5)* |
| II | 19 (24.1) | 2 (12.5) | 6 (9.5) | NA | 27 (17.1)* |
| III | 6 (6.3) | 2 (12.5) | 6 (9.5) | NA | 14 (8.2)* |
| IV | 33 (41.8) | 12 (75.0) | 42 (66.7) | NA | 87 (55.1)* |
| Missing | 2 (2.5) | 0 (0) | 3 (4.8) | NA | 46 (29.1)* |

**Supplementary Table 2**. Predictive value of CRP with outcome 'low' responder (vs high responder). Although CRP had an AUC of 0.71, at the clinical cut-off of 4 mg/L it had only a sensitivity of 30% and a specificity of 88% at baseline day 0. If used to identify patients that would benefit from adjuvant therapy, too many patients would be missed. An optimal cut-off of CRP to differentiate low from high responders was 1 mg/L that provided a sensitivity and specificity of 72% and 61%, respectively.

| Cut-Off | Sensitivity | Specificity | Positive likelihood ratio | Negative Likelihood ratio |
|---|---|---|---|---|
| 1 mg/L | 72% | 61% | 1.85 | 0.46 |
| 2 mg/L | 49% | 81% | 2.59 | 0.63 |
| 4 mg/L | 30% | 88% | 2.48 | 0.80 |
| 10 mg/L | 13% | 96% | 3.28 | 0.91 |
